# ECHOCARDIOGRAPHIC MANIFESTATIONS OF COVID 19 ILLNESS AND DEVELOPMENT OF PERSISTENT RV DYSFUNCTION AND PULMANARY HYPERTENSION AS A LONG TERM SEQUELAE OF COVID 19 ILLNESS: A STUDY AMONG PATIENTS OF SOUTH EAST ASIAN REGION

**DOI:** 10.1101/2023.05.26.23290622

**Authors:** Lalit Kumar, Himanshu Mahla, Neeraj Chaturvedi, Navdeep Singh Sidhu, Manisha Keshav, Shashi Mohan Sharma, Rajeev Bagarhatta, V.V. Agrawal, Vijay Pathak, Chandrabhan Meena, Deepak Maheshwari, Pradeep Meena, Balbir Singh Pachar, Sohan Kumar Sharma, Dinesh Gautam, Sarita Chaudhary, Dhananjay Shekhawat

## Abstract

**Objectives:** To study the Echocardiographic manifestations of covid 19 illness among patients admitted in our facility, Correlate MAPSE, TAPSE, PASP, CRP levels and CTSI among covid 19 patients with their 28 day outcome as survivors and non survivors and to look for evidence of residual RV dysfunction and Pulmonary hypertension using TTE after 1 year of follow-up.

**Study design:** Prospective observational study at various medical wards and ICUs in SMS medical college and associated hospitals.

**Methods:** 258 patients with a Covid-19 RT-PCR positive report from a throat or a nasal swab within 72 hours of admission were included in the study. Each patient underwent a complete clinical assessment and routine blood investigations including CRP levels were done. A complete transthoracic echocardiogram was done within 48 hours of admission. Patients also underwent a HRCT chest and CTSI scores were estimated. All patients were followed for a period of 28 days. The MAPSE, TAPSE, PASP, CTSI and CRP levels were then correlated with the outcome of the patient. The survivors again underwent a TTE at 1 year after their recovery from covid-19 illness to look for residual RV dysfunction by TAPSE and the development of pulmonary hypertension as measured by PASP using Bernoulli’s equation.

**Results:** Amongst patient of covid 19 illness the MAPSE, TAPSE, PASP, CTSI and CRP levels all correlated well with outcome of patients. While most covid-19 survivors recovered from their illness yet some patients showed evidence of persistent RV dysfunction and pulmonary hypertension even after 1 year of follow up.

## INTRODUCTION

The world in 2020 was devastated by the covid 19 pandemic which is now nearing towards its end. While most of the cases of covid 19 illness are mild and self-limiting, severe infection can lead to progressive pneumonia, multi-organ failure, and death.^1,2^Most patients with severe manifestation of covid 19 have some co existing comorbidity with cardiovascular disease being the most common. Up to 25% of patients with covid 19 illness develop some form cardiac involvement and this is associated with increased mortality among these patients.^2-6^ Covid-19 itself has been found to interact with and affect the cardiovascular system leading to myocardial damage and cardiac and endothelial dysfunction mainly via the Angiotensin-converting enzyme 2 (ACE-2) receptor.^7,8^Certain medications such as corticosteroids, antiviral medications, and immunological agents may have toxic side effects.^9-14^The extensive lung parenchymal involvement via mechanisms like oxidative stress, mitochondrial dysfunction, DNA damage, inflammation, hypoxia, endothelial dysfunction, and pulmonary micro-embolism can cause development of pulmonary hypertension or right ventricular dysfunction.^15^This type of PAH from covid 19 illness is thought to be a combination between type 3 and type 4 PAH. ^16^ Pagnesi et.al. estimated that PAH has a prevalence of 13% among covid 19 patients.^17^ however long term follow up studies of such patients are sparse. We therefore studied the echocardiographic parameters of heart among patients admitted to our facility with covid 19 illness, accordingly MAPSE, TAPSE and PASP were calculated and correlated with the initial CRP and CTSI of the patients. All these 4 parameters were then correlated with the outcome of patients as survivor or non-survivor at 4 weeks of illness. After 1 year the survivors were asked telephonically to come for a follow up visit and another TTE was done was done to look for evidence of residual RV dysfunction as measured by TAPSE or evidence of pulmonary hypertension by calculating PASP by Bernoulli equation.

## MATERIALS AND METHOD

The study was approved by Institutional Ethical Committee and a written informed consent was obtained from all patients prior to their inclusion in this study. Between May’2021 to sep’2021, 258 patients of covid -19 illness, admitted in various wards of SMS medical college and associated hospitals was enrolled in the study. Covid-19 was confirmed with throat or nasal swab RT PCR positive report. Patients enrolled into the study were subjected to complete history and clinical examination, all routine blood biochemistry including CRP along with Chest X-ray, ECG, and HRCT chest. The HRCT chest was used to calculate CTSI of the patient. A standard transthoracic echocardiogram was done in all patients within 48 hours of inclusion in the study. The MAPSE and TAPSE was calculated using M-Mode echocardiography at the lateral mitral and tricuspid valve annulus respectively, in the apical 4 chamber view. MAPSE value < 10 mm indicated LV dysfunction while TAPSE value <17 mm suggested RV dysfunction. The PASP was derived from TR jet velocity from apical 4 chamber view using the Bernoulli equation and adding estimated right atrial pressure from the IVC diameter and the respiratory variation in the IVC diameter in the standard subcoastal view. Patients with PASP ≥35 mm of Hg were considered having PAH. Patients were followed up for 28 days and then categorised as either survivors or Non survivors. The MAPSE, TAPSE and PASP were then correlated with CTSI as a marker of extent of lung parenchyma involvement, CRP as a maker of inflammation in the body and the outcome of patient as survivor or non-survivor at day 28. After 1 year the survivors were asked to come for a follow up visit telephonically and another TTE was done by the same operator to look for evidence of residual RV dysfunction as measured by TAPSE or evidence of pulmonary arterial hypertension using PASP calculated by Bernoulli equation and adding estimated right atrial pressure(PASP= 4TR^2^+RAP).

### INCLUSION CRIETERIA

patients of covid 19 illness, confirmed with a positive RT PCR report from throat or nasal swab with in last 72 hours were included in the study

### EXCLUSION CRIETERIA

Patients without a RT-PCR positive report, not willing to participate in study, admitted for >48 hours in the critical care, coronary artery disease(CABG/PTCA) or previous ventricular dysfunction, Known congenital heart disease, history of previous respiratory illness, pulmonary embolism, known cases of PAH and history using vasodilators like prostacyclin analogues, endothelin receptor antagonists, phosphodiesterase type 5 inhibitors, and guanylate cyclase stimulators were excluded from the study

### STATISTICAL ANALYSIS

The data was analysed using SPSS/22.0 software. The description of quantitative variables was performed using the mean, standard deviation (SD), median and quartiles. The correlation between variables was performed using the Pearson correlation coefficient, independent t test. A p value of <0.05 was considered significant.

## RESULTS

The study involved 258 patients with covid 19 illness of which 142(55.03%) were males and 116 (44.97%) were females. The mean age of the patients in the study was 49.53±13.43 years with a range of 18-79 years. The mean MAPSE, TAPSE, PASP, CRP, CTSI of the study population was were 13.8±2.08 mm, 16.54±3.44 mm, 26.12±19.3 mm of Hg, 43.39±46.35 mg/dl, 14.26±5.67 respectively. The 4 week in-hospital mortality rate among covid 19 patients was 28.69 % as 74 patients died within 28 days. The mortality rate was 25% in females as 29 females died while it was 31.69% among males as 45 males had died by 4 weeks. The average heart rate of patients with covid 19 was 102.91±22.26 beats per minute and 164 (63.56%) patients had tachycardia. Sinus tachycardia was the most common arrythmia seen in 142 (55.03%) patients, followed by atrial fibrillation seen in 13 (5.03%), and SVT in 7 (2.71%) patients. 2 patient (0.77%) had VT. Among bradyarrhythmia, 15 (5.81%) patients had bradycardia out of which 8 (3.1%) had sinus bradycardia, 2 (0.77%) had Mobitz type 1 av block while 5(1.93%) had CHB (TABLE 1).

**Table 1:** Baseline characteristics of patient

| STUDY VARIABLE | MEAN/FREQUENCY/RANGE |
| --- | --- |
| Age | 49.53±13.43 years (18-79 years) |
| Sex |  |
| Male | 142(55.03%) |
| Female | 116(44.97%) |
| Total (n) | 258 |
| Mean of various markers |  |
| CTSI | 14.26 ± 5.67 |
| TAPSE (in mm) | 16.54±3.44 |
| CRP (in mg/dl) | 43.39±46.35 |
| PASP (in mm of Hg) | 26.12±19.3 |
| MAPSE (in mm) | 13.8±2.08 |
| Prevalence of arrhythmias |  |
| Sinus tachycardia | 142(55.03%) |
| Atrial fibrillation | 13(5.03%) |
| SVT | 7(2.71%) |
| VT | 2 (0.77%) |
| Sinus bradycardia | 8(3.1%) |
| Mobitz type 1 AV block | 2 (0.77%) |
| CHB | 5 (1.94%) |
| Prevalence of RV dysfunction by TAPSE <17mm | 103(39.92%) |
| Prevalence of PAH (PASP=4 TR<sup>2</sup>+ RAP>35 mm of Hg) | 76(29.45%) |
| Prevalence of LV dysfunction by MAPSE < 10 mm | 43(16.66%) |
| OUTCOME at 4 weeks of illness |  |
| Survivors | 184(71.31%) |
| Non survivors | 74(28.69%) |
| Mortality rate |  |
| Females | 29(25%) |
| Males | 45 (31.69%) |

MAPSE was recorded as a measure of LV function and total 43(16.66%) of patients had a MAPSE of <10mm. The MAPSE of the patients correlated with both CTSI and CRP of the patient (p< 0.0001). The pearson correlation coefficient between MAPSE and CTSI was R= -0.5586, while it was R= -0.5215 between MAPSE and CRP. This suggested that negative correlation of MAPSE better with CTSI. The admission MAPSE correlated well with the day 28 outcome of patient (p<0.0001)

TAPSE was recorded as a measure of RV function and total 103 (39.92%) of patients had a TAPSE of <17mm. The TAPSE of the patient correlated with both CTSI and CRP of the patient(p<0.0001). The pearson correlation coefficient between TAPSE and CTSI was R= -0.6657, while it was R= -0.5913 between TAPSE and CRP suggesting that the negative correlation of TAPSE was more with CTSI. The admission TAPSE correlated well with the day 28 outcome of patient (p<0.0001)

TR jet velocity and the estimated right atrial pressure was measured to calculate PASP using the Bernoulli equation. In the study a total of 76 (29.45 %) of patients had PASP≥ 35 mm of Hg. The PASP of the patient correlated with both CTSI and CRP of the patient(p<0.008). The pearson correlation coefficient between PASP and CTSI was R= 0.612, while it was R= 0.596 between PASP and CRP suggesting PASP correlated better with CTSI. The admission PASP correlated well with the day 28 outcome of patient (p<0.007). Overall RV dysfunction was more common than Pulmonary arterial hypertension.

After 1 year the 184 survivors were contacted telephonically and 14 patients had died by 1 year, 27 patients could not be contacted, 41 patients refused for a follow up visit. In the remainder 102 patients another TTE was done to for evidence of residual RV dysfunction as measured by TAPSE or evidence of pulmonary hypertension by calculating PASP using Bernoulli equation and adding estimated right atrial pressure. Of the 102 patients 13(12.74%) had isolated TAPSE< 17 mm indicating RV dysfunction while isolated PASP ≥35 mm of Hg was found in 19(18.62%) patients indicating pulmonary arterial hypertension. 8(7.84%) patients had evidence of both RV dysfunction and pulmonary arterial hypertension. The mean MAPSE, TAPSE were less in non survivors while mean PASP, CRP and CTSI were higher amongst the non-survivor groups (TABLE 2 and GRAPH 1).

**Table 2:**
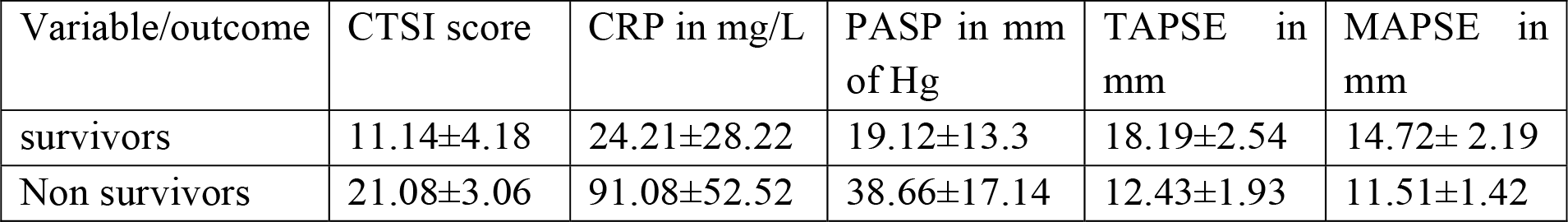
Mean CTSI, CRP and PASP And TAPSE score survivors and non survivors

**Table 3.** follow up data from Covid 19 survivors at 1 year follow up visit

|  |  |
| --- | --- |
| Total number of patient who followed up | 102 |
| Number of patient with TAPSE < 17 mm only<br>(RV dysfunction group) | 13 (12.74%) |
| Number of patients with PASP ≥35 mm of Hg only<br>(Pulmonary hypertension group) | 19(18.62%) |
| Number of patients with both TAPSE < 17 mm and PASP ≥35 mm of Hg<br>(Combined group) | 8(7.84%) |

The patient characteristics and study data are represented in the following tables.

**Figure 1.**
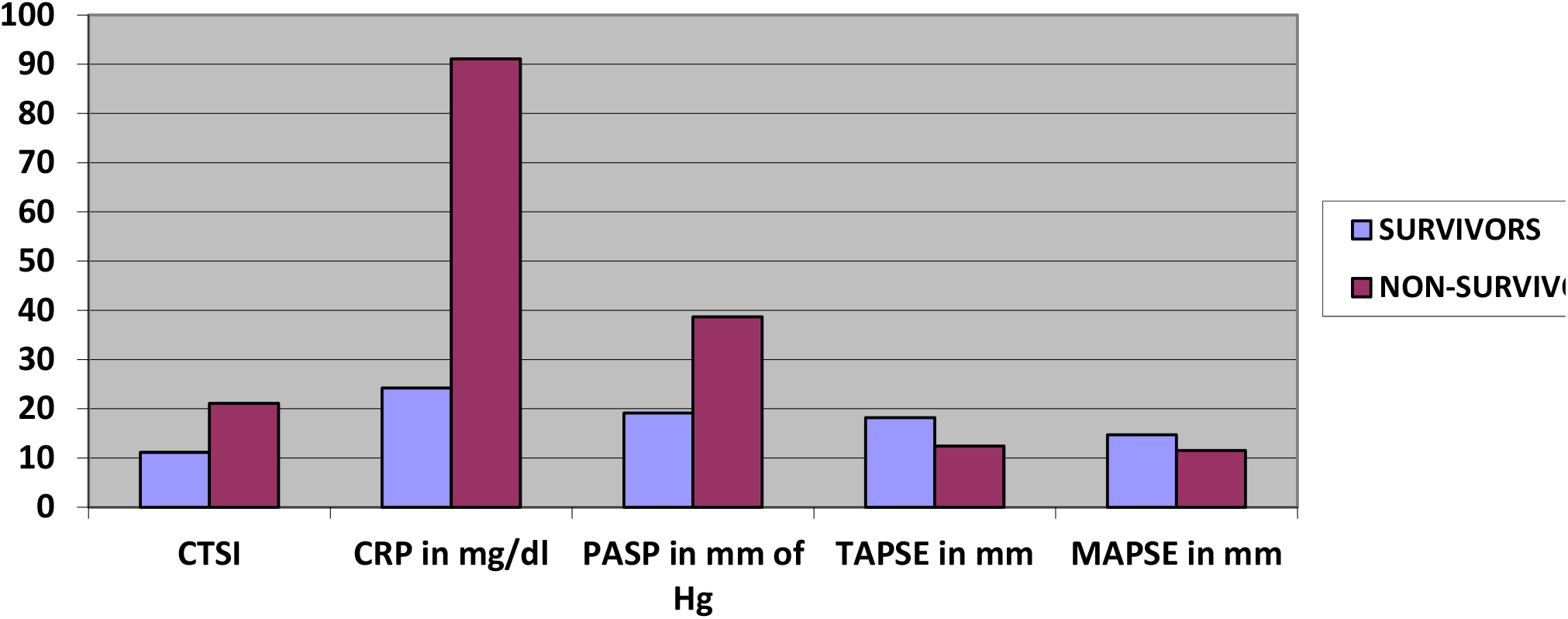
showing mean CTSI, CRP, MAPSE, TAPSE and PASP among day 28 survivors versus the non survivors

## DISCUSSION

Covid 19 illness is a complex illness and has a variable presentation including almost all organ systems of the body. The CVS and respiratory system is frequently involved in covid 19 illness and Multiple mechanisms have been suggested for their damage, these include 1) Cytokine Release Syndrome^11^ 2) Direct Myocardial Cell Injury^12^ 3) Acute Coronary Syndrome^13^ and 4) Other Possible Mechanisms include medications such as corticosteroids, antiviral medications, and immunological agents that have cardiotoxic side effects. The study involved 258 patients of covid 19 illness of which 142(55.03%) were males and 116 (44.97%) were females. The 4 week in-hospital mortality rate among covid 19 patients was 28.69 % as 74 patients died within the 28 days of illness. The mortality rate was 25% in females as 29 females died, while it was 31.69% among males as 45 males had died by 4 weeks. This data suggests that males have higher mortality rates in covid 19 illness.

When echocardiographic assessment of heart was done a total 103(39.92%) number of patients had a TAPSE of <17mm while PASP ≥35 mm of Hg was seen in 76(29.45%) number of patients. TAPSE is a measure of RV function while elevated PASP suggests pulmonary arterial hypertension. So, in covid 19 patients the RV dysfunction is more common than pulmonary hypertension. The presence of PAH and RV dysfunction is seen among many patients with ARDS who are treated in ICU. According to the Szekely Y et.al. in a study of 100 patients with covid 19 illness, the most common cardiac pathology was RV dilatation and dysfunction (observed in 39%of patients), followed by LV diastolic dysfunction (16%) and LV systolic dysfunction (10%).^18^ Similarly, Marc R Dweck et.al. studied a total of 1216 patients from 69 countries across six continents. Overall, 667 (55%) patients had an abnormal echocardiogram. Left and right ventricular abnormalities were reported in 479 (39%) and 397 (33%) patients, respectively, with evidence of new myocardial infarction in 36 (3%), myocarditis in 35 (3%), and takotsubo cardiomyopathy in 19 (2%).^19,20^ Previous studies have suggested that PAH is seen in 15% of patient with covid 19 illness. In our study the prevalence rates of RV dysfunction and PAH was high as ours was a sicker population as indicated by higher CTSI and CRP levels, also our prevalence rates overestimate the burden of both PAH and RV dysfunction in covid 19 illness, as most infections of covid 19 illness are subclinical or mild. It must be remembered that only the most serious patient got admission during the pandemic and were subsequentially evaluated in most studies.

When follow up TTE at 1 years was done most patient with covid 19 illness had recovered. However, 13(12.74%) patients had persistent RV dysfunction, 19(18.62%) had pulmonary arterial hypertension and 8(7.84%) had both RV dysfunction and pulmonary arterial hypertension. So PAH is more common than RV dysfunction in long term follow up, suggesting that the right ventricular recovers while because of extensive lung fibrosis pulmonary hypertension can set in. The development of RV dysfunction and pulmonary hypertension amongst the recovered patient can be explained by extensive pulmonary damage (due to interstitial and alveolar inflammation), alterations of pulmonary vasculature (induced by thrombotic/thromboembolic processes, endothelial injury, or, at least, hypoxic vasoconstriction), excessive use of positive end expiratory pressures used during admission. Although most patients with covid 19 illness recovered few patients had some residual RV dysfunction or pulmonary hypertension. Here it must be emphasized that since covid 19 virus infected millions of people around the world, even small percentage of covid 19 survivors with residual PAH would sum up to have a sufficient impact on the global prevalence of pulmonary hypertension.

## Conclusion

Covid 19 illness has frequent involvement of pulmonary parenchyma and myocardium. The 4-week mortality among hospitalized patients of covid 19 is 28.89%. The RV dysfunction assessed by TAPSE is seen in 39.92% of patients with covid 19 illness, while evidence of pulmonary hypertension when assessed by PASP using Bernoulli equation was seen in 29.45% of patients. When long term follow-up of covid 19 patients was done most of the patients had recovered but some patients still showed evidence of persistent pulmonary arterial hypertension and RV dysfunction. Even though these are small in numbers, because covid 19 virus led to millions of infections it may impact the global prevalence of pulmonary hypertension. This is especially true for countries that were hardest hit by the pandemic.

## Data Availability

ALL DATA IS AVAILABLE AND CONFEDENTIAL WITH THE AUTHOR

## REFRENCES

1. Bhatraju PK, Ghassemieh BJ, Nichols M, Kim R et al. Covid-19 in Critically Ill Patients in the Seattle Region - Case Series. N Engl J Med. March 30, 2020.

2. Zhou F, Yu T, Du R, Fan G et al. Clinical course and risk factors for mortality of adult inpatients with COVID-19 in Wuhan, China: a retrospective cohort study. Lancet. 2020; 395:1054-1062.

3. Guo T, Fan Y, Chen M, Wu X et.al. Cardiovascular Implications of Fatal Outcomes of Patients With Coronavirus Disease 2019(COVID-19). JAMA Cardiol. March 27, 2020.

4. Booth CM, Matukas LM, Tomlinson GA, Rachlis AR et al. Clinical features and short-term outcomes of 144 patients with SARS in the greater Toronto area. JAMA. (2003) 289:2801–9.

5. Guo T, Fan Y, Chen M, Wu X et al. Cardiovascular implications of fatal outcomes of patients with coronavirus disease 2019 (COVID-19). JAMA Cardiol. (2020) 5:811–8.

6. Liu PP, Blet A, Smyth D, Li H. The science underlying COVID-19: Implications for the cardiovascular system. Circulation. (2020) 142:68–78.

7. Wang D, Hu B, Hu C, Zhu F et.al. Clinical Characteristics of 138 Hospitalized Patients With 2019 Novel Coronavirus-Infected Pneumonia in Wuhan, China. JAMA. 2020 Mar 17;323(11):1061–1069.

8. Shi Y, Wang Y, Shao C, Huang J et.al. COVID-19 infection: the perspectives on immune responses. Cell Death Differ. 2020 May;27(5):1451–1454.

9. Xiong TY, Redwood S, Prendergast B, Chen M. Coronaviruses and the cardiovascular system: acute and long-term implications. Eur Heart J. 2020 May 14;41(19):1798–1800.

10. Dominguez-Erquicia P, Dobarro D, Raposeiras-Roubín S, Bastos-Fernandez G, Iñiguez-Romo A. Multivessel coronary thrombosis in a patient with COVID-19 pneumonia. Eur Heart J. 2020 Jun 07;41(22):2132

11. Lang RM, Badano LP, Mor-Avi V, Afilalo J et al. Recommendations for cardiac chamber quantification by echocardiography in adults: an update from the American Society of Echocardiography and the European Association of Cardiovascular Imaging. J Am Soc Echocardiogr. 2015; 28:1-39.e14.

12. Yancy CW, Jessup M, Bozkurt B, Butler J et al. 2013 ACCF/AHA Guideline for the Management of Heart Failure: Executive Summary: A Report of the American College of CardiologyFoundation/American Heart Association Task Force on Practice Guidelines. Circulation. 2013; 128:1810–1852.

13. P. Goyal, J.J. Choi, L.C. Pinheiro, et al. Clinical characteristics of Covid-19 in New York City N Engl J Med: 382 (2020), pg. 2372–2374

14. Gawalko M et.al. COVID-19 associated atrial fibrillation: incidence, putative mechanisms and potential clinical implications. Int J Cardiol Heart Vasc. 2020; 30: 100631

15. Potus, F.; Mai, V.; Lebret, M.; Malenfant, S.; Breton-Gagnon, E.; Lajoie, A.C.; Boucherat, O.; Bonnet, S.; Provencher, S. Review: The pathophysiology of COVID-19 and SARS-CoV-2 infection novel insights on the pulmonary vascular consequences of COVID-19

16. Van Dongen, C.; Janssen, M.; van der Horst, R.; van Kraaij, D.; Peeters, R.; van den Toom, L.; Mostard, R. Unusually rapid development of pulmonary hypertension and right ventricular failure after COVID-19 pneumonia. EJCRiM 2020, 7, 001784

17. Pagnesi M, Baldetti L, Beneduce A, Calvo F, Gramegna M, Pazzanese V, Ingallina G, Napolano A, Finazzi R, Ruggeri A, Ajello S, Melisurgo G, Camici PG, Scarpellini P, Tresoldi M, Landoni G, Ciceri F, Scandroglio AM, Agricola E, Cappelletti AM. Pulmonary hypertension and right ventricular involvement in hospitalised patients with COVID-19. Heart. 2020 Sep;106(17):1324–1331

18. Szekely Y, Lichter Y, Taieb P, Banai A, Hochstadt A, Merdler I, Gal Oz A, Rothschild E, Baruch G, Peri Y, Arbel Y, Topilsky Y. Spectrum of Cardiac Manifestations in COVID-19: A Systematic Echocardiographic Study. Circulation. 2020 Jul 28;142(4):342–353

19. Marc R Dweck, Anda Bularga, Rebecca T Hahn, Rong Bing et.al. Global evaluation of echocardiography in patients with COVID-19, European Heart Journal - Cardiovascular Imaging, Volume 21, Issue 9, September 2020, Pages 949–958

20. Perazzo H et.al.; RECOVER-SUS Brasil Group. In-hospital mortality and severe outcomes after hospital discharge due to COVID-19: A prospective multicenter study from Brazil. Lancet Reg Health Am. 2022 Jul;11:100244.

